# *“Everything created by a white man is for pagans”:* Understanding the Barriers to Childhood Immunization in North-Eastern and North-Western Nigeria

**DOI:** 10.1101/2024.03.29.24305068

**Authors:** Eno-Obong Eti, Angela Odiachi, Leanne Dougherty, Matthew Alabi, Adetayo Adetunji, Adebola Adedimeji

## Abstract

**Background:** Despite investments by the Nigerian government and international organizations in childhood immunization to combat child mortality, coverage in many northern states remains below the national average, thereby increasing the risk of vaccine-preventable diseases. This study describes the barriers to immunization in six states in Northeast and Northwest Nigeria, which have the lowest vaccination coverage rates in the country.

**Method:** The socioecological framework informed the design of focus group discussions that we conducted with 93 caregivers and 91 community influencers from March to July 2022 to understand their perceptions of the barriers to immunization uptake. We thematically analyzed data that were inductively and deductively coded in NVivo version 12 software.

**Results:** Supply-side service delivery challenges were the main systems-level barriers to vaccine uptake. These included poor provider attitude, shortage of health workers, vaccine stock-out, long wait-times, and inequitable service delivery. Other barriers were, rumors, misconceptions, vaccine distrust, socio-cultural and religious norms at the community level, and spousal and family disapproval at the interpersonal level. Limited autonomy and mobility, poor vaccine knowledge and awareness of immunization benefits, and concerns about side effects were important individual-level barriers.

**Conclusion:** Strategies for improving immunization uptake should include intensifying community sensitization, improving the numbers and quality of the health workforce, involving men and traditional institutions through context-appropriate channels, and addressing socio-cultural norms and concerns about adverse events after immunization

## Introduction

Immunization is among the most effective and cost-efficient public health interventions for reducing child morbidity and mortality (1–3). For decades, vaccines have safely reduced the burden of polio, measles, and smallpox, and allowed 2 - 3 million children annually to grow up healthy and happy. Between 2010 and 2018, the measles vaccine alone prevented 23 million deaths (4,5). In spite of this, vaccination coverage has stagnated in recent years and dropped since 2020. Globally, an estimated 25 million children under 1 year did not receive basic vaccines (list) in 2021. This represents six million more children than in 2019, and a decrease in coverage from 86% to 81%. Of these 25 million children, one-quarter lived in Nigeria (6,7).

The Nigerian government and international organizations have taken considerable measures to combat rising child mortality rates in Nigeria, particularly through interventions aimed at increasing routine immunization (RI) coverage (8). Despite these efforts, millions of children remain at risk of death from vaccine-preventable diseases. In 2019, Nigeria topped the World Health Organization’s list of countries with the highest number of under-5 deaths (9,10). The 2021 Multiple Indicator Cluster Survey (MICS) showed that 1 in 10 children died before their fifth birthday, and 64 per cent of children 12 - 23 months did not receive all required vaccines (7). These rates were much higher in northern Nigeria with 27 per cent unvaccinated children in the Northwest and Northeast, compared to 5 – 11% (11,12) in other zones.

Immunization in northern Nigeria remains a sensitive subject due to pervasive misconceptions, myths, and false information, resulting in low community demand and vaccine-uptake (13). Thus, the persistence of vaccine-preventable diseases in northern Nigeria has remained a public health concern (14). Several factors responsible for observed low child immunization rates include caregivers’ lack of knowledge, and/or disapproval, (15,16). myths about vaccines, fear of side effects, maternal education, socioeconomic status, and a lack of trust in health personnel (15,17-19). At the health system level, weak supply chains, poor delivery of services, scarce human resources, funding gaps, poor resource accountability, and weak governance have been cited as potential barriers to uptake (20).

The Bill and Melinda Gates Foundation (BMGF) and Aliko Dangote Foundation (ADF) have partnered with the governments of six northern Nigeria states (Kano, Kaduna, Bauchi, Borno, Sokoto, and Yobe) since 2012, through a memorandum of understanding (MoU) to strengthen routine immunization systems and address funding challenges. The program aimed to enhance collaboration to improve vaccine supply chain and logistics, enhance service delivery, address human resource capacity, and boost community trust and demand for RI and primary healthcare (PHC) services in these partner states.

This study, nested in a larger mixed-method evaluation of the RI MoU approach, explored the barriers associated with childhood immunization uptake in the six MoU states. The purpose is to assess and better understand the challenges that remain despite the huge investments aimed at improving vaccination coverage. Understanding these barriers would provide useful information for improving RI programs and inform immunization policies, especially given Nigeria’s commitment to the Global Vaccine Action Plan’s goal (GVAP (Reference).

## Materials and Methods

### Study Area and Design

This qualitative study was conducted in Bauchi, Borno, Yobe in the Northeast, and Kaduna, Kano, and Sokoto in the Northwest) states of Nigeria. These six states were the focal states of the MoU program. Vaccination coverage rates across these states are low at 5.8% (Sokoto), 13.6% (Borno), 16.5% (Bauchi), 23.0% (Kano), 34.5% (Kaduna), and 49.2% (Yobe) in 2021) (7). The Social-Ecological Model (SEM) which underpinned this study, examined the multiple levels of influence on immunization behavior. It posits that behavior both influences and is influenced by diverse contexts (21). This model considers interactions between individuals and their social networks, community structures, and institutional/systems/policy contexts. Using this framework, we developed a focus group discussion guide to elicit information from community leaders and other stakeholders on how these multiple levels constituted barriers to RI uptake.

### Study Population and Sampling

Using purposive sampling, we selected two categories of participants namely, caregivers and community leaders/influencers. Caregivers were women of reproductive age (15 – 49 years), who were mothers of under-2 children and partial or non-adopters of immunization services. Partial adopters were mothers who had initially taken up but did not complete immunization for their children, while nonadopters were mothers who did not access immunization services for their children. Community influencers comprised traditional, religious, women, and youth leaders, and traditional birth attendants (TBAs) who lived in study communities and worked in collaboration with the State Primary Healthcare Development Agency (SPHCDA) on maternal, neonatal and child health (MNCH) issues in these communities. Study participants were recruited with the assistance of health facility workers. These service providers identified potential respondents from health facility defaulter tracking registers and from line-listing records of community leaders/influencers who worked with the health facilities. Line-lists contained key information about all newborns in the communities. Community leaders used these lists to track all children who were eligible for immunization. The study took place in 2 to 4 local government areas (LGAs) per study state. The selection of these LGAs was based on the distance from the state capital, and security considerations.

### Data Collection

A total of 24 FGDs were conducted (12 with partial and non-adopters of routine immunization services, and 12 with community leaders/influencers). All study instruments were developed in English and translated into Hausa (the local language in the states). All FGD guides were pre-tested in a location outside the study communities. Feedback from the pilot testing informed modifications to the FGD guides. Fieldwork was implemented simultaneously in the six states between March and April 2022. Each state data collection team comprised seven experienced and trained qualitative interviewers who were familiar with the subject matter and study states. The FGDs were conducted face-to-face at primary healthcare facilities within each community and lasted about 75 minutes each. Data gathering activities were conducted in ethically and culturally sensitive environments. Interviews were facilitated by two researchers: one interviewer and one notetaker. All interviews were digitally recorded, stored on password-protected computers, and transcribed verbatim using a transcription protocol developed by the research team.

### Data Analysis

All interviews conducted in Hausa were translated into English. Transcribed interviews were then transferred to NVivo version 12 software and coded by a team of eight trained coders. We adopted a hybrid deductive-inductive thematic analysis approach; by first identifying themes from the FGD guides and study objectives, and later exploring and adding emergent patterns and themes in the data to our analytical codebook. For rigor, the research team initially coded the same transcripts, to ensure that the codes were consistently applied, and discrepancies were resolved through consensus.

### Ethical Considerations

Ethical approval was granted by the National Health Research Ethics Committee (NHREC/01/01/2007-17/01/2022), all six states’ ethical committees (Bauchi (NREC/03/11/19B/2021/10); Borno (073/2021); Kaduna (MOH/ADM/744/VOL.1/1171); Kano (SHREC/2022/3078); Sokoto (SKHREC/016/2022) and Yobe (MOH/GEN/747), and the Population Council’s Institutional Review Board in New York (Protocol number 992). Written informed consent was also provided by all study participants before commencing interviews. Each participant provided individual written informed consent prior to the interviews, and for the FGD to be audio-recorded.

## Results

### Barriers to Immunization Uptake

#### Individual Level Barriers

##### Limited female autonomy and decision-making power

Women’s limited autonomy and decision-making power contributed to low immunization uptake. When mothers encountered opposition from husbands, parents and sometimes in-laws, they were unlikely to take their children for immunization, regardless of their personal views.

> I am really in support of it [immunization] at first for my three children. But on the third child, his dad [husband] said he was tired of the injection; he did not understand what was going on. You would buy drugs with money, but when you to the hospital they give the injection for free. He said we should just stop going for it. But I did not stop going until I finished collecting five doses for the third child. And for the fourth child… he didn’t support me. He stopped me and said I should not go again. –**Partial-adopter, Borno state**.

##### Adverse events after immunization

Concerns about side effects of vaccinations also influenced caregivers’ decision to continue immunization. Participants who reported their children suffered from side effects such as fever and malaise, particularly after receiving pentavalent vaccines. Although some caregivers understood that these side effects were transient and common, many reported that this resulted in their refusal to continue. In some cases, caregivers were not counselled by service providers regarding these side effects, thus leading many to refuse the continuation and completion of required doses.

> The reason why he [husband] stopped me was the children got fever when we returned [after immunization]. When he [husband] asked why and I told him that I collected an injection for them, he asked, “Why?” And that I should not go again. That on the quest of getting drugs for catarrh, I have brought something new upon the child. –**Partial-adopter, Borno state**.
>
> We have not gotten any information concerning the importance of immunization. Not from community meetings or mosques, not at all. - **Non-adopter, Yobe state**.

#### Interpersonal Level Barriers

##### Spousal permission to access health services

Interpersonal factors that hindered vaccine uptake included pressure from caregivers’ social networks: peers, spouses, family members, and social support networks-that often-discouraged caregivers from seeking immunization services. Reports that husbands and other family members restricted women from taking their children for immunization were common. Such disapproval emanated from mistrust of vaccines, concerns about potential side effects and adverse events following immunization (AEFI), sociocultural norms, and limited knowledge of the advantages of immunization.

> Honestly, I was discouraged by my husband… We recorded a small complication [side effect] in the child and since then he [husband] has restricted me from going for another session. He feels the immunization poisons the child. –**Partial-adopter, Kano state**.

#### Community Level Barriers

Community-level barriers contribute to a dis-enabling environment in which childhood immunization is discouraged. This includes social and religious norms; rumors and misconceptions; and mistrust of vaccines.

##### Misconceptions and myths about vaccines

For instance, myths regarding conventional medicine were the most frequently cited community barrier to immunization uptake. Caregivers described concerns that vaccines cause infertility, contribute to the onset of other illnesses, and sometimes death of the child.

##### Religious beliefs and norms

Similarly, caregivers and community influencers in Kano and Yobe states extensively discussed concerns based on religious beliefs. Some community members believed that vaccination was forbidden by their religion or that seeking care from health facilities was contrary to religious teachings. Community leaders in Kano reported that many community members believed the ability to heal or be disease-free was an act of God that could only be obtained through prayer.

> There are those whose religion prevents them from accessing RI. Some of them stay in urban areas but they rejected polio vaccines, and some of them believe that “Everything created by a white man is for pagans.” We hear them say these things, and we live with them. Some of them see coming to health facilities as forbidden. –**Male Community Influencer, Yobe state**.

##### Health provider-patient gender discordance

Concerns about the gender of health providers also influenced immunization uptake, since women were often the parent/caregivers who brought children to health facilities for immunization. Community influencers revealed that some men were hesitant to allow their wives to visit health facilities where the providers were male, thereby, reflecting deeply entrenched cultural norms about other men having close interactions, as is often the case in health service provision, with their spouse, even in the health facility setting. This in turn hindered access of women and children to health services, including immunization.

> Most of our hospitals don’t have women. And [according to] our tradition in the village, by the time they say all the staffs are males, some people will not allow their wives to go there. – **Female Community Influencer, Sokoto state**.

Fears surrounding vaccine safety and rumors about the motivations of community influencers in facilitating demand also influenced community members’ perception of immunization. Many assumed that these influencers were more concerned with the financial gain they “must” have received from the government for facilitating service use, than the community’s health.

> Sometimes we face a lot of problems. You will direct someone to go [for vaccination]. They will go behind you and insult you. For instance, I went to tell some people to go and collect this injection, but they refused. I insisted that they must collect it and went there with them. After collecting the vaccine, they got back to their houses and started gossiping that they won’t be surprised that I have collected money. That is why I am clamoring for people to go and collect it [vaccine]. – **Female Community Influencer, Borno state**.

In addition, some participants questioned the underlying governments and donors’ motivations in providing free vaccines to healthy children when medical care for sick children was not free, raising concerns about potential hidden agenda.

> Since the day that the announcers came to tell people to go and collect vaccine injections, he [husband] said, “Why would they say that people should go and collect it free of charge, since they do not give drugs free of charge when the child is taken to the hospital at the time the child is sick?” They should help with drugs when the child is sick, not when he is healthy. – **Non-adopter, Borno state**

Surprisingly, this sentiment was shared by some community influencers, even though they were better informed about the importance of routine immunization so they can educate and create awareness among community members.

> There was a time I brought my two children to the health facility, and it happened that free drugs were recently brought to the health facility by the government. They refused to give me [the free drugs], and I complained… I promised not to take my children to a health facility again after that incident. What stops them from giving us free services? …Why do they offer free services only when they come to our houses [for immunization]? – **Female Community Influencer, Yobe state**.

#### Systems Level Barriers

##### Poor health worker attitude

Previous unpleasant experiences with service providers were discussed as the major institutional barrier to immunization uptake. Many caregivers of children who were eligible for immunization reported feeling disrespected by health workers, particularly during previous antenatal and delivery visits. Such absence of respectful care prompted caregivers to seek care in other distant facilities to avoid future mistreatment. However, transport costs to such alternate facilities were high, which, in turn, hindered caregivers’ ability to access care.

> I ran away from [the facility] because they don’t value their patients, they don’t treat us right, in every way. I went there to give birth and was humiliated. Since then, I discourage people from going there to access care. – **Non-adopter, Sokoto state**.

##### Ineffective documentation of immunization appointments

Some caregivers reported forgetting scheduled immunization dates because health workers only verbally informed them of upcoming appointments and did not record these dates on vaccine appointment cards. Further, when mothers missed appointment dates, they ultimately decided to forego immunization for fear of possible reprimand from healthcare providers.

They [health workers] honestly don’t write the time you should return. They would just tell you to return after four weeks. You could sometimes forget to return. – **Partial-adopter, Borno state**.

##### Distance to health facilities

The consensus among all study participants that availability of and accessibility to health facilities impacted service utilization. Many emphasized the inadequacy of health facilities in meeting the population’s needs. Similar reports emerged in discussions with community influencers, who disclosed challenges on linking some women to routine immunization services due to a lack of health facilities in their communities and the consequent financial burden of seeking care in distant facilities.

##### Inadequate human resource capacity at health facilities

This was also identified as an system level barrier to service uptake, resulting in long wait times for immunization at health facilities. For example, participants reported that it was common at many facilities for only one service provider to be solely responsible for vaccine retrieval from the local government cold stores, administering vaccines (and providing other health services), and vaccine record-keeping.. Service delays were further compounded by facility personnel prioritizing, sometimes inequitably, who received care, causing some caregivers to return home without immunizing their children when the facility ran out of vaccine supplies.

> The challenges in our clinic are the workers, the work is too much for one person… Sometimes when you go, she [service provider] will be doing the immunization and still writing reports and sometimes they will be looking for her at the center [to undertake other health tasks]. So that is a challenge. – **Partial-adopter, Kaduna state**.
>
> Sometimes you get there early but they attend to others who come after you. That was why I stopped going. - **Partial-adopter, Sokoto state**.

##### Vaccine supply shortages and stock-outs

Another systems barrier was vaccine stock-outs, particularly in Yobe and Bauchi states. Caregivers reported visiting health facilities multiple times without their children receiving vaccines due to vaccine stock-outs. Some women highlighted that returning home with an unvaccinated child after seeking permission from their spouse to visit the facility undermined their credibility and engendered suspicion in their households.

> Some days you come [visit the facility], and it [vaccine] is finished. And because you have told your husband you were going for immunization, coming back to tell him you didn’t get the immunization makes him feel like you were lying, and he will not allow you to go back the following week. You see we are not on our own, we are under him so we can’t go until he gives us permission. - **Partial-adopter**, Yobe **state**.

*Lack of incentives* was an additional challenge reported. Community influencers suggest that incentives were a motivating factor for immunization uptake. Typically, state governments provide mosquito nets to caregivers who bring their children to health facilities for immunization. However, some states have reported a stock-out of nets in the months preceding data collection. Without incentives, community influencers reported difficulty in persuading mothers to immunize their children.

> We used to give out mosquito nets when we administer measles and yellow fever vaccines. Now it’s been about 6 months since we ran out of stock and that is discouraging some of the mothers from coming. – **Female Community Influencer, Kaduna state**.

## Discussion

Our findings show that the attitude of health workers was an important factor in the uptake of routine immunization services. Mothers were reportedly reluctant to visit facilities due to previous negative experiences with service providers and fear of reprimand for missing scheduled immunization appointments. Tarrant & Gregory (2023) reported in their Canada study that reprimands from providers frequently upset mothers and left them feeling inadequate about their ability to care for their children, resulting in future refusals to visit health facilities (22). This highlights the importance of respectful care and positive relationships between clients and service providers in facilitating vaccine uptake.

Poor documentation of vaccine appointment dates was also noted as a systems-level barrier in the six states. Due to improper documentation of appointment dates, many caregivers missed vaccination appointments. Given that most women were already burdened with child and home care, immunization cards could have served as a reminder and a vital health record for tracking immunization status. These findings are consistent with the 2021 National Immunization Coverage Survey (NICS) which found low immunization to be directly related to low vaccination card availability (7). Health workers’ failure to complete immunization cards raises capacity concerns and may indicate an overworked workforce.

The inability to and delays in accessing health services were also key barriers identified in this study. Many non-adopters cited long distance to a health facility as a barrier to service uptake. Several studies have documented that geographic accessibility to health facilities offering routine immunization services was a major determinant of immunization coverage in many areas of northern Nigeria (23-25). It is however worth noting that all focus group discussions for this study were held at health facilities within these communities, suggesting that the non-prioritization of immunization services by caregivers may be an additional underlying cause. More so because our study discovered that for some caregivers, immunization was seen as unnecessary for healthy children.

Discrimination in terms of the inequitable prioritization of who received services was also raised by women in Sokoto state and has been shown to be a barrier to service uptake (26). Additionally, long waiting time was also a persistent issue in the states. In this study, long waiting time was directly linked to staffing shortages at health facilities. These shortages could also be the reason for poor documentation of vaccine appointment dates and improper counseling on the side effects of immunization. Furthermore, incentives, such as free mosquito nets were found to be a motivation for immunization uptake. Despite the substantial evidence demonstrating the positive relationship between incentives and immunization uptake, a study conducted in Sokoto state found that immunization uptake may decrease to previous low levels or lower once incentives are withdrawn (19). Our study supports this finding, as we discovered that in some states, caregivers stopped immunizing their children when incentives were no longer provided. This raises sustainability concerns and potential unintended consequences regarding incentives. The persistent demand for incentives appears to be driven by the need for communities to feel that they are receiving tangible benefits from immunization.

A major objective of the RI PHC MoU is to strengthen vaccine supply and availability. However, our findings suggest the persistence of vaccine stockouts in certain states, particularly in Yobe and Bauchi. During focus group discussions, caregivers described returning home with unvaccinated children due to stockout. This raised suspicion from spouses, and mistrust in households. In a highly patriarchal and conservative region where women’s mobility is often restricted, returning home without an immunized child gave husbands the impression that immunization was being exploited as an excuse to visit other places. This poses a challenge to spousal permission for future immunization of children and echoes other studies on the need to engage men through appropriate channels including traditional institutions (27).

Overall, study participants cited rumors and misconceptions, socio-cultural and religious norms, and mistrust as the most common social and cultural barriers to immunization. Several studies have documented that the increased number of polio campaigns in Nigeria was viewed to be suspicious by some populations (28-30). The content of these documented rumors was similar to those reported in our study with some caregivers expressing the belief that vaccinations are clandestine family planning tools that cause infertility, death, or serve as a means of population control. Importantly, our findings also reveal that key opinion leaders may contribute to these misconceptions, as some questioned the government’s motivations for providing free vaccines while other health services for children were not free. This raises concerns about the knowledge and capacity of the community influencers, who anchor immunization sensitization in communities. If influential opinion leaders hold such views, it is plausible that false information about immunization could spread within communities.

We also found that caregivers in Kano and Yobe were often conflicted between their religious beliefs and the utilization of immunization services. Many believed that it was religiously prohibited to visit a health facility as prayer was the only real protection against disease. Others were convinced that orthodox medicine or any invention by the West was for “pagans.” These findings were similar to those from a study conducted in Sokoto, which identified a reliance on faith for protection against disease as one of the barriers to vaccine uptake (13).

Our findings highlight that the absence of appropriate AEFI counselling and management fueled vaccine-related myths leading to service discontinuation. A major challenge was the accompanying side effects of vaccines, particularly with pentavalent vaccines. Some caregivers were hesitant to return for future vaccine doses when their children experienced any side effects, especially fever. This may explain the low coverage of Penta 3 in some northern Nigeria states, which is as low as 11% in states such as Sokoto, a far cry from the Global Vaccine Action Plan’s goal of 90% (12). This could be responsible for the suspicions surrounding free vaccines in the communities. Other studies in northern Nigeria have reported similar findings (13, 31, 32). This finding also calls for interventions to ensure healthcare workers can provide adequate AEFI counselling on adverse outcomes following vaccination and emphasizes the need for continuous capacity building for service providers.

Limited knowledge and awareness on immunization was also noted from conversations with caregivers. Some women claimed to have never received any information on vaccines or its benefits. Nonetheless, knowledge of immunization benefits did not always translate to the utilization of the service. Many mothers who were aware of the benefits of immunization were unable to immunize their children due to opposition from their spouses or other influential family members. Further interactions with women revealed that this was often attributed to women’s limited autonomy and mobility. When women were confronted with resistance from their husbands, they preferred to avoid utilizing immunization services for fear of the consequences of disobedience. Often, the reason for these refusals was the lack of female health workers at health facilities to provide services to women. Female healthcare providers play a crucial role in providing acceptable health services for female clients, especially in the context of conservative northern Nigeria. Consequently, proactive management of this workforce could provide opportunities to address these barriers.

### Limitations

This study only explores barriers to immunization from caregivers’ and community influencers’ perspectives; the views of service providers are not represented. Thus, the findings reported here do not effectively highlight system (supply-side) challenges to routine immunization uptake. Our study did not include male participants/fathers of young children, who are key decision makers. Our findings present only the views of female caregivers. Despite this, this study adds to the body of knowledge on barriers to immunization uptake in northern Nigeria and provides useful information to policy makers and program implementers to improve routine immunization uptake.

## Conclusion

This study’s findings reveal that the factors that influence the utilization of immunization services are complex. Most of the barriers to childhood immunization found in the study include societal norms, rumors, poor quality of health services, gender dynamics, and non-prioritization of immunization, all of which point to longstanding barriers to service utilization in northern Nigeria. Immunization programs would need to focus on intensifying efforts to promote increased sensitization, particularly regarding myths and misconceptions, and increasing male involvement activities tailored to spouses and fathers, who play a crucial role in determining whether a child is immunized. Sociocultural and other gender-related norms that function as barriers to service adoption must also be addressed through social and behavioral change initiatives. There is a need for immunization programs to strengthen training, counselling, and reporting of adverse events by service providers. Many mothers have mentioned side effects as a primary factor for their refusal to start or continue immunization of their children. Their spouses did not allow them to also take up services for this same reason. If health workers provide adequate counselling on side effects, such women are likely to continue with subsequent vaccinations. Furthermore, while it is necessary to continue working with community influencers to create demand for routine immunization, it is also essential to strengthen efforts towards capacity building for these groups. As community influencers are a key source of information in society, it is pertinent that they have the correct messaging to avoid spreading misinformation.

## Data Availability

All data relevant to the publication can be obtained upon reasonable request to the Population Council.

## Acknowledgement

The authors appreciate all study participants across all six states where the study was conducted and the local research assistants who undertook data collection activities. We also specially thank the State Primary Health Care Development Agency in all six states for their support during the research work.

